# Biologic DMARD Access and Medication Cost-related Nonadherence in Rheumatology Patients in Canada: A Cross-sectional Survey

**DOI:** 10.1101/2023.01.20.23284839

**Authors:** Anne Marie Holbrook, Avrilynn Ding, Sue Troyan, Sandra Costa, Mark Matsos, Michael R. Law, Selena Gong, Apurva Dixit, Gary Foster, Nader Khalidi

## Abstract

**Background:** Cost-related nonadherence to prescription medications affects many Canadians and is associated with negative self-perceptions of health. Biologic disease modifying anti-arthritic drugs (bDMARDs) are costly drugs recommended for certain patients with rheumatoid or psoriatic arthritis and ankylosing spondylitis. We investigated access and cost-related nonadherence (CRN) to bDMARDs compared to other therapies for such patients in Ontario.

**Methods:** We conducted a cross-sectional telephone survey of adult patients recruited from two academic rheumatology practices in Hamilton, Ontario, asking demographic and socioeconomic characteristics, drug plan coverage, medication cost-related cutbacks, opinions on the value of bDMARDs, and assistance with costs from health professionals. CRN was defined by patient self-report of not using or using less than prescribed amount of medication, due to cost.

**Results:** 104 patients (mean age (SD) 61(12) years) participated, including 77 (74%) women, 57 (54.8%) taking bDMARDs, and 27 (25.9%) with household income <$40,000 annually. CRN was reported by 19 (18.3%) participants with no significant difference between those taking versus not taking bDMARDs (risk difference (95% CI): -0.10 (−0.25, 0.04); p=0.19). 37 (64.9%) of those taking bDMARDs reported that they would not take them if they had to pay the full cost. Overall, few patients reported that they would ask their doctor (17.3%) or pharmacist (15.4%) for help with reducing prescription costs.

**Conclusion:** CRN prevalence was relatively high amongst these rheumatology patients despite access to public and private funding mechanisms. Patients expressed a reluctance to ask their doctor or pharmacist for help in reducing their medication costs.

## Introduction

Adherence to chronic medication internationally is notoriously poor, on average at approximately 50% after six to 12 months, with 30% of patients not even picking up the prescription (Anderson et al., 2020; Ho et al., 2009; Osterberg & Blaschke, 2005). The cost of prescription medications is thought to be a barrier for many patients to access the healthcare that they need (Tang et al., 2014). Cost-related nonadherence (CRN) is defined as medication underuse because of cost; this includes unfilled prescriptions, delayed prescriptions, taking smaller doses or less frequent doses than prescribed (Law et al., 2018). Not only is poor adherence associated with more adverse outcomes for patients, it also unnecessarily inflates health care spending because of wastage (Heisler et al., 2010; Kennedy & Morgan, 2009). In Canada, CRN to medications range from 5.1% to 10.2%, with factors predicting CRN including high out-of-pocket spending, low income or financial flexibility, lack of drug insurance, younger age, and poorer health (Holbrook et al., 2021).

Rheumatoid arthritis (RA), psoriatic arthritis (PsA) and ankylosing spondylitis (AS) are the more common inflammatory arthritides and amenable to biologic disease modifying anti-rheumatic drugs (bDMARDs). Each disease has other treatment options including nonsteroidal anti-inflammatory drugs (NSAIDs), corticosteroids, conventional DMARDs (cDMARDs) (Gossec et al., 2016; Rohekar et al., 2015; Singh et al., 2016). Systematic reviews do not show superiority of bDMARDs compared to combinations of cDMARDs for RA, but are preferred to cDMARDs for severe PsA and for patients with AS where NSAIDs provide insufficient benefit (Saad et al., 2008; Wu et al., 2015).

Since biologic drugs are now the single most costly group of medications for public drug plans in Canada, their judicious use is required (CIHI, 2019). Most bDMARDs are covered by Ontario’s public drug benefit plan (OPDP) through a process called ‘frequently requested exceptional access’ which requires communication by prescribers to OPDP as to how the individual patient meets specific access criteria (Ministry of Health, 2020). The criteria for public plan reimbursement involve in each case the documentation of failure of two previous conventional therapies or combinations, and renewal requires documentation of improvement with bDMARD use (Ministry of Health, 2020). Even with access restricted to only these situations approaching cost-effectiveness, the bDMARDS are estimated to cost approximately $300 million annually for rheumatologic conditions in Ontario (Tadrous M et al., January 2020. DOI 10.31027/ODPRN.2020.01).

Our objective in this study was to investigate bDMARD access, adherence and opinions regarding value and costs, for patients with rheumatoid arthritis, ankylosing spondylosis, and psoriatic arthritis cared for in academic rheumatology practices.

## Methods

We designed a cross-sectional survey study using a questionnaire delivered by trained research staff by telephone. The study was approved by Hamilton Integrated Research Ethics Board (HiREB #11-077). Reporting followed the guidelines of the STROBE adaptation for cross-sectional surveys (Equator Network, 2020; von Elm E).

The setting was the practices of two full-time rheumatology specialists in Hamilton, Ontario, Canada. These practices draw referrals from a regional catchment of approximately two million people. Both specialists reported having detailed knowledge of bDMARD drug plan funding processes.

Inclusion criteria were adults age 18 years or older with one of Rheumatoid Arthritis (RA), Psoriatic Arthritis (PsA) or Ankylosing Spondylitis (AS). Exclusion criteria included patients not fluent in English with no accessible translator, too frail, or significant cognitive impairment. Potentially eligible patients were first screened for presence of exclusion criteria by their rheumatologist, who sent an information letter about the study by mail to the remaining group, asking them to reply if interested in participating. Recruitment ended in 2016.

Once informed consent was obtained, the questionnaire itself took 15 to 30 minutes to complete. We asked about demographic and socioeconomic characteristics (education level, annual household income), prescription medication used for arthritis, drug plan coverage (co-pay, deductibles, caps, etc) for prescription medications, out-of-pocket expenditure for prescriptions and other medications, cost-related medical and nonmedical cutbacks, opinions on the value for money of bDMARDs, and requests for assistance with costs from health professionals.

Questions related to cost-related nonadherence were adapted from our previous survey on CRN (Holbrook A, 2020; Zheng B, 2012) which were subsequently adapted for the Canadian Community Health Survey (Law et al., 2012; Law et al., 2018). Participants were asked about leaving prescriptions unfilled, delaying picking up a prescription, not filling a prescription, skipping doses, lowering doses, or substituting prescribed medication with a cheaper alternative.

The main analyses were descriptive. CRN was based on the 6 questions described above, each with scoring options of never=zero, rarely=one, sometimes=two, often=three, and always=four. Total CRN scores were calculated for each participant, with highest possible score of 24. CRN was defined as a response of at least “sometimes” to one or more of the six questions (e.g., total score two or more). For modeling, we used a method to estimate risk differences (Spiegelman & Hertzmark, 2005), to be able to characterize absolute differences between groups. Univariate modeling was used to examine age, income, insurance coverage, medication status, co-payment and out-of-pocket expenses (defined as the amount that the patient paid for prescription medication that was not reimbursed) as possible predictors of CRN as a dichotomized variable (total CRN ≥ two versus < two, Multivariable modeling was planned if there were sufficient numbers in each score category. Data analyses were performed using SAS v9.4 (SAS Institute, Inc., Cary, North Carolina).

## Results

Of approximately 400 patients who met inclusion criteria and were mailed invitation letters to participate by their rheumatologist, 104 patients participated in the study. Detailed participant characteristics are provided in Table 1. 77 (74.0%) were female, mean age was 60.8 years old (standard deviation [SD] 12.1), 62 (59.6%) had at least some post-secondary school education and 27 (25.6%) reported household income as less than $40,000 annually. Drug coverage included a mix of private and public drug plans, with only four (3.8%) participants reporting entirely self-pay. 57 (54.8%) participants were taking a bDMARD, compared to 47 (45.2%) not taking a bDMARD. Patients receiving bDMARDs were more likely to be female (80.7% vs 66.0%) and under 65 years of age (75.4% vs 38.3%). A majority of respondents (88 (84.6%) paid $100 or less per month out-of-pocket for all prescription medications. This included 50/57 (87.7%) patients using bDMARDS and 38/47 (80.9%) patients not using bDMARDs. No patient taking bDMARDs paid the full out-of-pocket cost for their medication.

**Table 1.** Baseline Characteristics

| **Characteristics** | **All Patients**<br>**(n=104)** | **Patients taking**<br>**bDMARDS**<br>**(n=57)** | **Patients not**<br>**taking**<br>**bDMARDS**<br>**(n=47)** |
| --- | --- | --- | --- |
| **Age, yr** , mean (SD)<br>(Minimum, Maximum) | 61 (12)<br>(24, 86) | 56 (12)<br>(24, 78) | 66 (10)<br>(41, 86) |
| **Sex**<br>Male†<br>Female | 27 (26)<br>77 (74) | 11 (19)<br>46 (81) | 16 (34)<br>31 (66) |
| **Highest Education**<br>Less than high school<br>Graduated high school<br>Some post-secondary<br>Graduated university<br>Post-graduate | 12 (12)<br>30 (29)<br>35 (34)<br>16 (15)<br>11 (11) | 6 (11)<br>12 (21)<br>23 (40)<br>12 (21)<br>4 (7) | 6 (13)<br>18 (38)<br>12 (26)<br>4 (9)<br>7 (15) |
| **Household Income** (\$/yr)<br>Less than 20,000<br>20,000 - 39,999<br>40,000 - 69,999<br>70,000 – 100,000<br>Greater than 100,000<br>Refused/No response | 7 (6.7)<br>20 (19.2)<br>33 (31.7)<br>15 (14.4)<br>24 (23.1)<br>5 (4.8) | 4 (7)<br>15 (26)<br>14 (25)<br>10 (18)<br>13 (23)<br>1 (2) | 3 (6)<br>5 (11)<br>19 (40)<br>5 (11)<br>11 (23)<br>4 (9) |
| **Out-of-Pocket Rx Expense** (\$/mo)<br>0-19<br>20-100<br>101-200<br>201-300<br>>300<br>Refused/No response | 52 (50.0)<br>36 (34.6)<br>9 (8.7)<br>1 (1.0)<br>2 (1.9)<br>4 (3.8) | 34 (59.6)<br>16 (28.1)<br>4 (7.0)<br>1 (1.8)<br>1 (1.8)<br>1 (1.8) | 18 (38.3)<br>20 (42.6)<br>5 (10.6)<br>0 (0)<br>1 (2.1)<br>3 (6.4) |
| **Drug Insurance Coverage**§<br>Only private coverage<br>Only government coverage<br>Government and private<br>Full Out-of-Pocket<br>Unknown | 47 (45.2)<br>29 (27.9)<br>23 (22.1)<br>4 (3.8)<br>1 (1) | 32 (56.1)<br>14 (24.6)<br>10 (17.5)<br>0 (0)<br>1 (1.8) | 15 (31.9)<br>15 (31.9)<br>13 (27.7)<br>4 (8.5)<br>0 (0) |

† Data cells are number (%) unless otherwise specified
§ Drug insurance coverage referred to the arthritis drugs only.

Nineteen of 104 (18.3%) participants including 13/57 (22.8%) taking bDMARDs and 6/47 (12.8%) not taking bDMARDs met our definition of CRN (a score of two or more on one or more domain), chi-square p = 0.187. Univariable analyses identified predictors of CRN as age (6.9% increase in likelihood of CRN for every 10 year decrease in age, p=0.010), out-of-pocket expenditure of $100 or more compared to < $20 per year, p=0.007, and income of < $40,000 compared to ≥ $40,000 per year, p=0.035 (details in Table 3).

More patients taking bDMARDs compared to patients not taking bDMARDs (13 (22.8%) vs six (12.8%), p = 0.21) used one or more strategies to cover the cost of medication, most commonly cutting back on leisure and recreation items, followed by clothing and food (Table 2). 46 (80.7%) patients taking bDMARDs reported that the bDMARD effectiveness and safety was worth their cost, but 37 (64.9%) also reported that they would not be taking bDMARDs if they had to pay the full cost themselves. Those who were prescribed biologics were more likely to report that their physicians asked how much they paid for drugs and were more likely to consider the cost of their medications, compared to those not prescribed a biologic, 47 (82.4%) versus 22 (46.8%), and 37(64.9%) versus 24 (51.1%) respectively. However, only a small number of patients, 18 (17.3%) overall, 14 (24.6%) taking bDMARDs versus four (8.5%) not taking bDMARDs, asked for assistance with drug costs from their physician, and 16 (15.3%) overall, 12 (21.1%) using bDMARDs versus four (8.5) not taking bDMARDs, from their pharmacist.

**Table 2.**
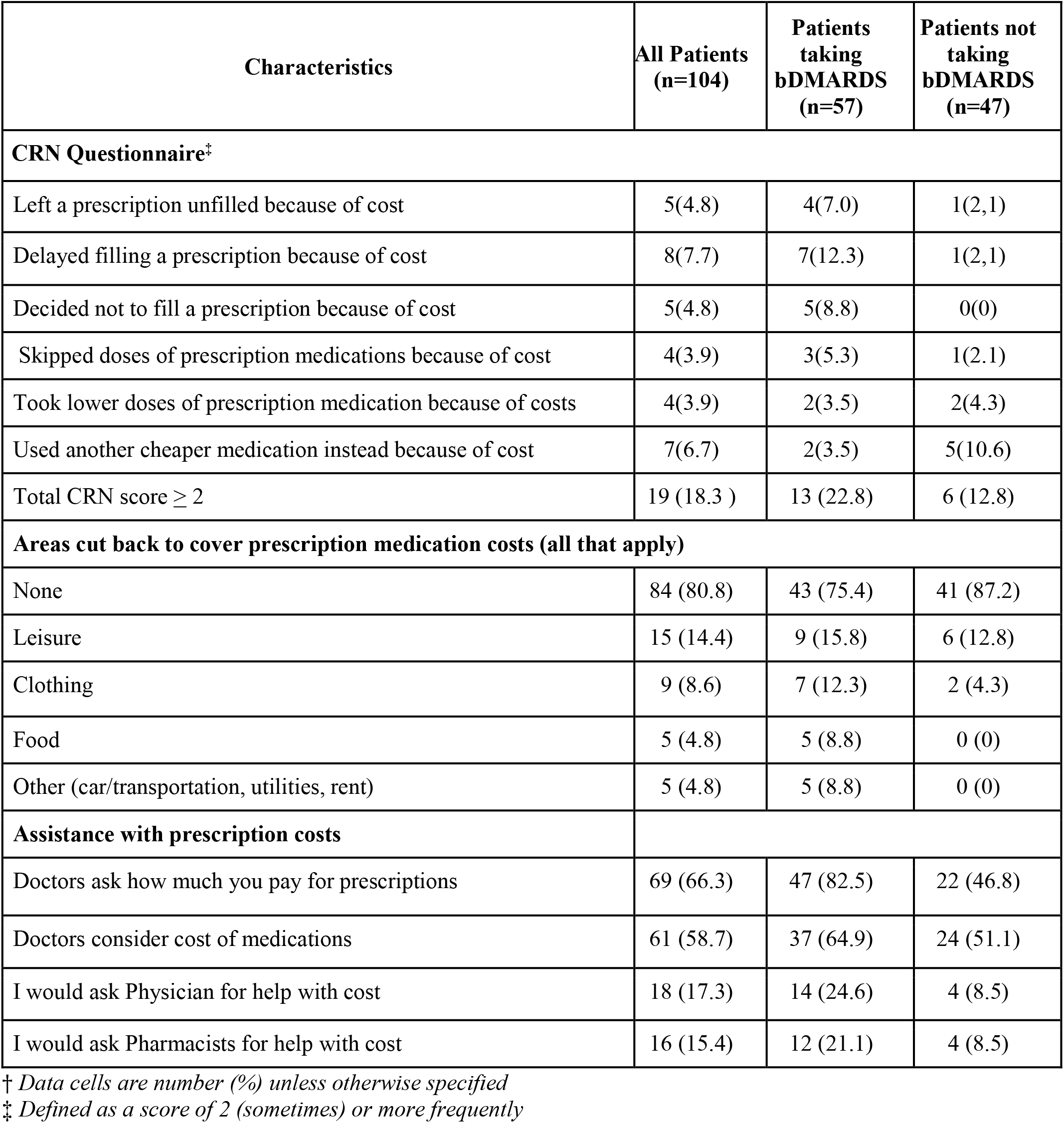
Cost-related Non-adherence Results^†^.

**Table 3.** Univariable Analyses of Predictors of CRN Total Score.

| **Variable** | **Comparison** | **Risk Difference (95 CI)** | **p-value** |
| --- | --- | --- | --- |
| **Age** | Increase in 10 years | -0.069 (-0.121, -0.017) | 0.010 |
| **Household Income (\$/yr)** | < \$40,000 vs ≥ \$40,000 | 0.208 ( 0.015, 0.402) | 0.035 |
| | < \$70,000 vs ≥ \$70,000 | 0.089(-0.059, 0.236) | 0.241 |
| **Sex** | Male vs Female | -0.097 (-0.246, 0.053) | 0.204 |
| **Biologics use** | Yes vs No | 0.100 (-0.044, 0.245) | 0.174 |
| **Out-of-pocket expense (\$/mo)** | \$100+ vs <\$20 | 0.404 ( 0.110, 0.698) | 0.007 |
| | \$20-\$99 vs <\$20 | 0.071 (-0.075, 0.216) | 0.343 |

## Discussion

Our cross-sectional examination of access to expensive bDMARDs for their most common rheumatologic indications for patients in academic practices, revealed that the self-reported physician awareness of public drug plan funding criteria and requirements for bDMARDs was used regularly to gain access for patients. While the prevalence of cost-related nonadherence in this group of patients was higher than the 9.6% average for Canadians receiving prescriptions, there was no significant difference between patients on biologics and those not taking biologics (Heidari et al., 2018). Although most patients taking bDMARDs felt that their benefit was worth the cost, only a minority would continue taking them if paying the full cost.

A systematic review of costs and medication adherence for patients with RA found that rates of adherence ranged from 18.2% to 48% at one year post-initiation, and out-of-pocket costs were the main predictors (Heidari et al., 2018). These studies included all medications for arthritis. Given that bDMARDs are approximately five times more expensive than cDMARD combinations, CRN is likely to be more common for these patients. Several studies suggest that 1-year rates of nonadherence to bDMARDs in international cohorts with inflammatory arthritides, are high at 20% to 54%,(Dalén et al., 2016; Stolshek et al., 2018) but most do not address cost-related nonadherence specifically. A survey of older adults with RA in Medicare reported CRN in 18.4%, a rate three and a half times those with other chronic diseases, and these patients were two and a half times more likely to spend less on basic necessities (Harrold et al., 2013). Although bDMARDs are in the treatment algorithms of RA, PsA and AS, their expense has modified their place in therapy (Mian et al., 2019). A review of patient attitudes and opinions showed some ambivalence towards the drugs, with a sense of privilege involved in access, but concern because of their cost and their perceived toxicity which was influenced by the experience of patients’ acquaintances rather than by the summary evidence (Kelly et al., 2018).

Our study is limited by a small sample size, a single urban setting and restriction to academic practices, and lack of details on medications for those patients not taking bDMARDs, making it difficult to generalize to other types of settings.

The implications of this study are several. Clinicians need to regularly check on patients’ medication adherence and drug insurance coverage, and offer assistance in reducing medication costs. Policy makers in Ontario may be reassured that these patients can get access to bDMARDs but should be concerned by the high rate of CRN.

Despite good physician advocacy leading to high levels of access to public and private plan coverage for all types of bDMARDs, CRN prevalence was still relatively high amongst these patients and did not differ between those taking versus not taking bDMARDs. This study should be replicated in a larger sample of patients.

## Data Availability

All data produced in the present work are contained in the manuscript.

## Acknowledgments and Affiliations

The study was funded through a Canadian Institute of Health Research grant titled “Cost related Non-adherence to prescription drugs; A Multi-Method Study” (PI, Michael Law).

